# Structured Outpatient Specialty Care and Faster Posttraumatic Stress Disorder Symptom Improvement: A Matched Cohort Study

**DOI:** 10.64898/2026.02.27.26347276

**Authors:** Sara Khor, Hannah Klempner, Emily R. Dworkin, Andrew Schwehm, Mill Brown, Adam Chekroud, Matt Hawrilenko

## Abstract

**Objective:** Although trauma-focused psychotherapies are effective for posttraumatic stress disorder (PTSD), recovery under routine outpatient conditions remains variable. This study examined whether a structured Specialty Care (SpC) model integrating condition-focused clinicians, higher session frequency, and proactive care navigation was associated with faster PTSD symptom improvement than standard outpatient care.

**Methods:** A retrospective matched cohort study (2023–2025) was conducted among U.S. adults with elevated PTSD symptoms (PTSD Checklist for DSM-5 [PCL-5] ≥31) in an employer-sponsored digital mental health platform. Access to SpC was determined by employer benefit. Propensity-score matching balanced cohorts on baseline severity and demographics. Outcomes included PCL-5 trajectories and time to symptom-defined recovery; depressive symptoms (PHQ-9) were secondary. Linear mixed-effects and proportional hazards models were applied.

**Results:** The matched sample included 356 SpC and 9,409 standard outpatient care participants. SpC participants received higher psychotherapy dose and faster early follow-up, showed steeper PCL-5 decline (β = −1.3 per log-week, p< .001; between-group difference, 3.8 points, d = 0.30), and higher likelihood of symptom-defined recovery (HR, 1.31; 95% CI, 1.10–1.57; 29% vs. 23% at 12 weeks). Post hoc analyses indicated the effect was largely attributable to greater and earlier therapy dose. Depressive symptoms improved in both groups without significant differences in categorical recovery.

**Conclusions:** A structured outpatient specialty care model was associated with modestly faster symptom-defined PTSD recovery by delivering more psychotherapy sooner. Redesigning outpatient care delivery may be a practical lever for improving routine PTSD outcomes without requiring a novel psychotherapy protocol.

**Highlights:**

- A structured outpatient delivery model with higher frequency and earlier therapy and more intensive care support was associated with modestly faster symptom-defined PTSD recovery than standard outpatient care.
- The benefit appeared largely attributable to delivering more psychotherapy sooner.
- Restructuring outpatient care to front-load session intensity and proactive engagement may improve real-world PTSD outcomes.

## Introduction

Posttraumatic stress disorder (PTSD) affects approximately 4.7% of U.S. civilian adults annually and is associated with substantial functional impairment, workplace disruption, and increased healthcare utilization.^1–3^ Although evidence-based trauma-focused psychotherapies are effective, recovery in routine outpatient settings remains variable. One contributor may be the organization of care: even when evidence-based treatments are available, patients may experience delays in treatment initiation, widely spaced sessions, limited care coordination, or difficulty remaining engaged during periods of symptom exacerbation.

Routine outpatient PTSD care is commonly delivered in generalist psychotherapy settings, where treatment frequency, follow-up cadence, and use of trauma-focused protocols may vary by clinician, patient preference, acuity, and service availability. For patients with elevated PTSD symptoms, low-frequency or patient-paced care may be poorly aligned with clinical need, particularly when avoidance, comorbidity, safety concerns, or social stressors interfere with treatment engagement and may contribute to premature dropout.^4,5^ More intensive outpatient and residential programs may address some of these needs, but are costly, difficult to access, and disruptive to work and family responsibilities.^6^

Several delivery components have been associated with better engagement or faster symptom reduction, including condition-focused treatment protocols, greater session frequency, and coordinated care navigation.^7–13^ However, these components are usually evaluated separately or in controlled trials. Less is known about whether integrating them within a structured outpatient pathway is associated with faster PTSD symptom improvement under routine conditions.

The Specialty Care (SpC) pathway was designed to improve outpatient care delivery for patients with higher-acuity behavioral health conditions, including trauma, eating disorders, substance use disorders (SUD), and severe mood or anxiety symptoms. SpC was implemented within an employer-sponsored digital mental health platform that also offered standard outpatient behavioral health services through a Core Program. The pathway included condition-focused clinician assignment, higher-frequency psychotherapy, proactive outreach, and ongoing care navigation.

The objective of this study was to evaluate whether enrollment in this structured SpC pathway was associated with differences in psychotherapy utilization, PTSD symptom trajectories, and time to symptom-based clinical improvement compared with the Core Program. We hypothesized that SpC enrollment would be associated with more timely early follow-up, greater cumulative psychotherapy dose, and faster improvement in PTSD symptoms. This study evaluates SpC as a service-delivery pathway, not as an evaluation of specific PTSD psychotherapies.

## Methods

### Study Design and Data Source

We conducted a retrospective cohort study using longitudinal clinical and utilization data from an employer-sponsored digital mental health benefit platform serving approximately 443 U.S. employers across diverse industries. The dataset included structured symptom, demographic, and utilization records. The Institutional Review Board deemed the study exempt as not human subjects research; informed consent was not required. This study followed the Strengthening the Reporting of Observational Studies in Epidemiology (STROBE) guideline for cohort studies.^14^

### Measurement-Based Care and Program Access

The platform employs measurement-based care, with validated self-reported symptom assessments administered at enrollment and every 2 to 4 weeks via email prompts, in-session, or before sessions in the virtual waiting room. Assessment completion was voluntary, and clinicians were financially incentivized to incorporate measurement-based care. Access to SpC was determined by employer benefit design. All participants had access to the Core Program. The comparison group comprised Core participants whose employers did not purchase SpC and were therefore structurally unable to enroll, reducing individual-level selection in SpC. However, employer-level differences in benefit selection may remain a source of residual confounding. In both programs, participants had access to 6–12 employer-sponsored psychotherapy sessions, with additional sessions via health plan coverage. Sponsored sessions could also be used for psychiatric medication management. Care navigation and a 24/7 crisis line were available.

### Care Models

The Core Program provided outpatient behavioral health care through care navigators, licensed therapists, and medication managers, including psychotherapy, medication management when indicated, measurement-based care, care navigation, and platform-based telehealth and scheduling. Core participants predominantly self-scheduled psychotherapy visits through the platform and could select clinicians with or without documented trauma-focused training. Care navigation in the Core Program focused on access facilitation and coordination and was typically brief and front-loaded around intake.

SpC was a structured outpatient delivery pathway comprising four condition-specific tracks: SUD, eating disorders, severe mood/anxiety disorders, and trauma-related symptoms. When multiple needs were present, hierarchical triage prioritized stabilization of higher-acuity conditions. SpC differed from the Core Program through condition-focused clinician assignment, higher-frequency psychotherapy, and more intensive care navigation. Members were assigned to clinicians with condition-focused training and/or certification in manualized protocols, with all trauma-pathway clinicians required to have training in at least one of Prolonged Exposure, Cognitive Processing Therapy, Eye Movement Desensitization and Reprocessing, Accelerated Resolution Therapy or trauma-focused Cognitive Behavioral Therapy. Psychotherapy could occur up to twice weekly when clinically indicated. Care navigation began with an approximately 45-minute proactive triage intake to support psychotherapy scheduling and early engagement and continued with condition-specific outreach 1–2 times per month, acuity-based monitoring, engagement support, and escalation when indicated.

Both programs used the same digital infrastructure, including telehealth, scheduling, secure messaging, symptom assessments, self-guided tools, and between-session engagement features. Use of specific digital features, between-session engagement, session-level protocol use, treatment fidelity, and protocol completion were unavailable. Accordingly, we evaluate SpC as a service-delivery pathway and not as a test of specific psychotherapy protocols. eTable 1 summarizes key differences between the Core Program and SpC pathway and indicates which components were observed.

### Study Population

Eligible participants were U.S. adults with elevated PTSD symptoms, defined by baseline PTSD Checklist for DSM-5 (PCL-5) score ≥31,^15^ between January 1, 2023, and December 31, 2025, who also completed a baseline Patient Health Questionnaire-9 (PHQ-9) (eFigure 1). For SpC participants, baseline was the PCL-5 assessment closest to enrollment (prior to or within 90 days after enrollment); for Core participants, baseline was the first recorded PCL-5. Analyses were limited to assessments within half a year of baseline. The primary PCL-5 cohort required a baseline PCL-5 ≥31 and at least one follow-up PCL-5. The PHQ-9 cohort required baseline PHQ-9 ≥10 and ≥1 follow-up PHQ-9. Baseline characteristics were compared by follow-up status to evaluate potential selective attrition.

### Measures

The primary outcome was PTSD symptom severity measured by the PCL-5, a 20-item self-report measure assessing PTSD symptoms over the past month. Because trauma exposure and clinician-diagnosed PTSD were unavailable, thresholds are symptom-defined proxies. We defined symptom-defined recovery as PCL-5 <31;^15^ low-symptom status as PCL-5 ≤20;^16^ clinically meaningful improvement as a ≥10-point reduction from baseline;^17^ and a composite of clinically meaningful improvement plus recovery. Secondary depressive-symptom outcomes (PHQ-9) used analogous thresholds: recovery <10, low-symptom status <5, and clinically meaningful improvement ≥5-point reduction.^18,19^ Service-utilization measures included psychotherapy initiation, cumulative psychotherapy sessions, time from first to second session, care navigation visits, and medication management visits.

### Statistical Analysis

Baseline clinical severity did not differ by follow-up status, although individuals without follow-up tended to have greater social determinants of health (SDOH) needs (eText 1). Propensity scores, estimated separately for the PCL-5 and PHQ-9 cohorts, modeled the probability of SpC enrollment based on baseline PCL-5, PHQ-9, and GAD-7 severity, SUD risk, suicide risk, self-reported drugs or alcohol concerns, age, self-reported gender, self-reported race and ethnicity, and number of documented SDOH needs. Race and ethnicity were socially constructed demographic covariates for matching and not for race- or ethnicity-specific inference. Each SpC participant was matched to multiple Core participants with similar propensity scores using optimal variable-ratio matching (matchit R package) to improve covariate balance while preserving the full analytic sample. Balance was assessed using weighted standardized mean differences, with values below 0.10 indicating acceptable balance.

Cumulative psychotherapy, navigation, and medication visits within 12 weeks of treatment initiation were estimated with weighted mean cumulative functions (Nelson-Aalen) using inverse probability weights and cluster-robust standard errors. Early follow-up was assessed by time to the second psychotherapy session (Kaplan-Meier and weighted Cox models, censored at 14 days) and cumulative sessions within 14 days, a window selected to capture early treatment cadence while minimizing the influence of later step-down or benefit-limit effects.

Symptom trajectories were estimated with linear mixed-effects models (REML), with fixed effects for log-weeks since baseline, SpC, and their interaction. Random intercepts were included for participants and matched stratum. These models provide unbiased estimates under a missing-at-random assumption and include all matched participants regardless of follow-up assessment count. The PCL-5 model adjusted for baseline PHQ-9 and high SUD risk, and the PHQ-9 model adjusted for baseline high SUD risk.

Time-to-event analyses used interval-censored proportional hazards models to evaluate the association between SpC enrollment and time from baseline to each symptom threshold. Because symptom assessments occurred intermittently, threshold attainment was assumed to occur between the last assessment above the threshold and the first assessment meeting it. Participants who did not meet a threshold were right censored at their last available assessment. Models included SpC enrollment, baseline covariates, and a patient-specific latent severity term derived from symptom trajectory models to account for baseline differences in symptom burden. Robust variance estimates accounted for matching.

### Sensitivity Analyses

Sensitivity analyses restricted the sample to participants initiating psychotherapy within 30 days of baseline and examined whether effects differed between SpC participants assigned to the trauma vs. SUD pathways. Eating disorder and severe mood/anxiety pathways were excluded from subgroup analyses because of insufficient sample sizes.

Post hoc exploratory analyses examined whether cumulative psychotherapy and care navigation exposure accounted for differences in PTSD symptom trajectories associated with SpC enrollment. Mixed-effects models analogous to the primary analysis sequentially added cumulative psychotherapy and navigation contacts completed before each assessment. These analyses were exploratory.

## Results

### Cohort Characteristics and Matching Balance

A total of 9,770 members met initial inclusion criteria for the PCL-5 analysis (eFigure 1). Following propensity score matching, the analytic sample included 356 SpC participants from 57 employers and 9,409 Core Program participants from 386 employers, with excellent covariate balance (all standardized mean differences <0.10; Table 1; eFigure 2). Mean age was 38 years (SD 12); mean baseline PCL-5 and PHQ-9 scores were 52 (SD 14) and 15 (SD 6), respectively. Median time to last follow-up assessment was 61 days (SpC) and 56 days (Core). A separately matched PHQ-9 cohort (296 SpC; 8,014 Core) also achieved balance (eFigure 2).

**Table 1.**
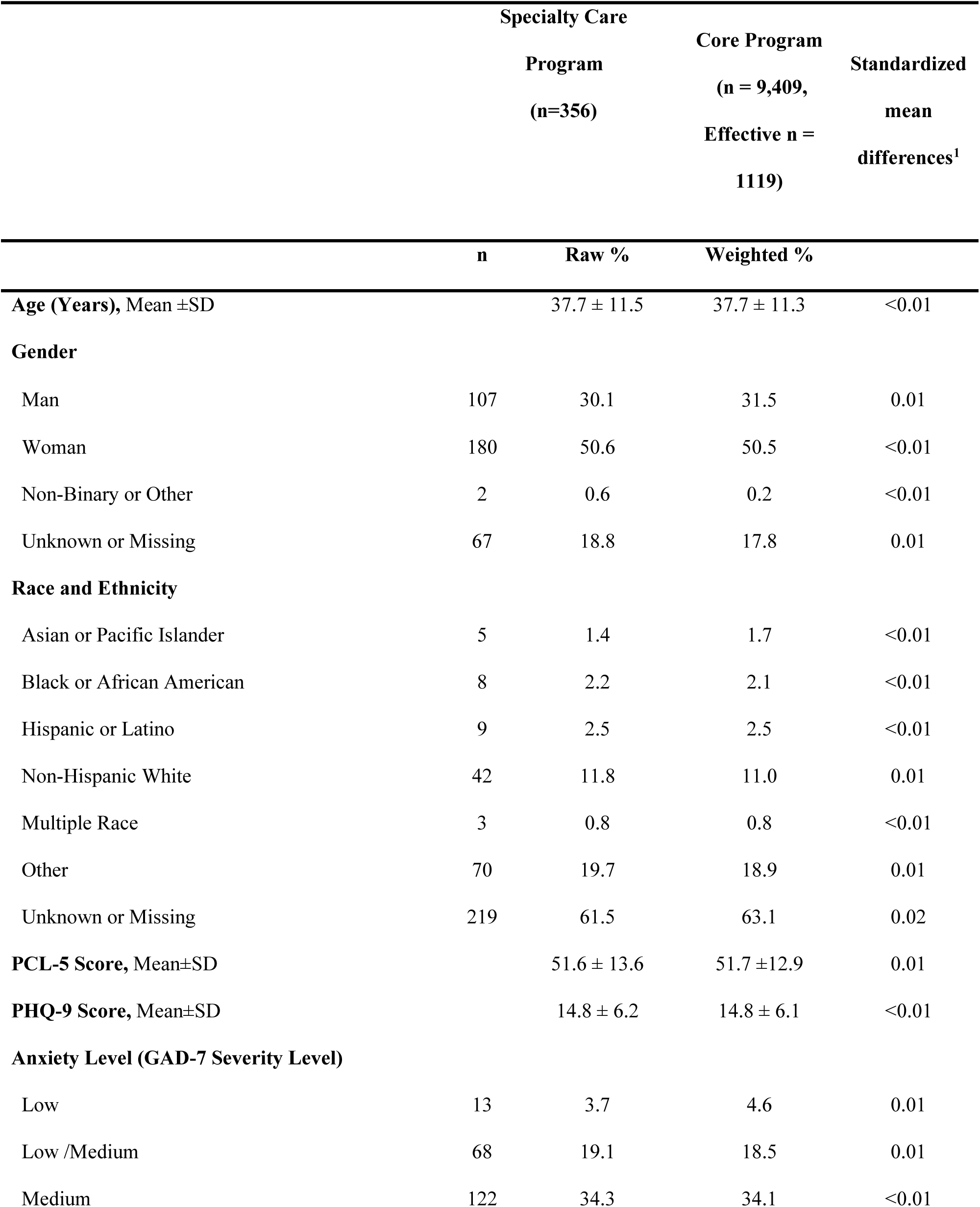
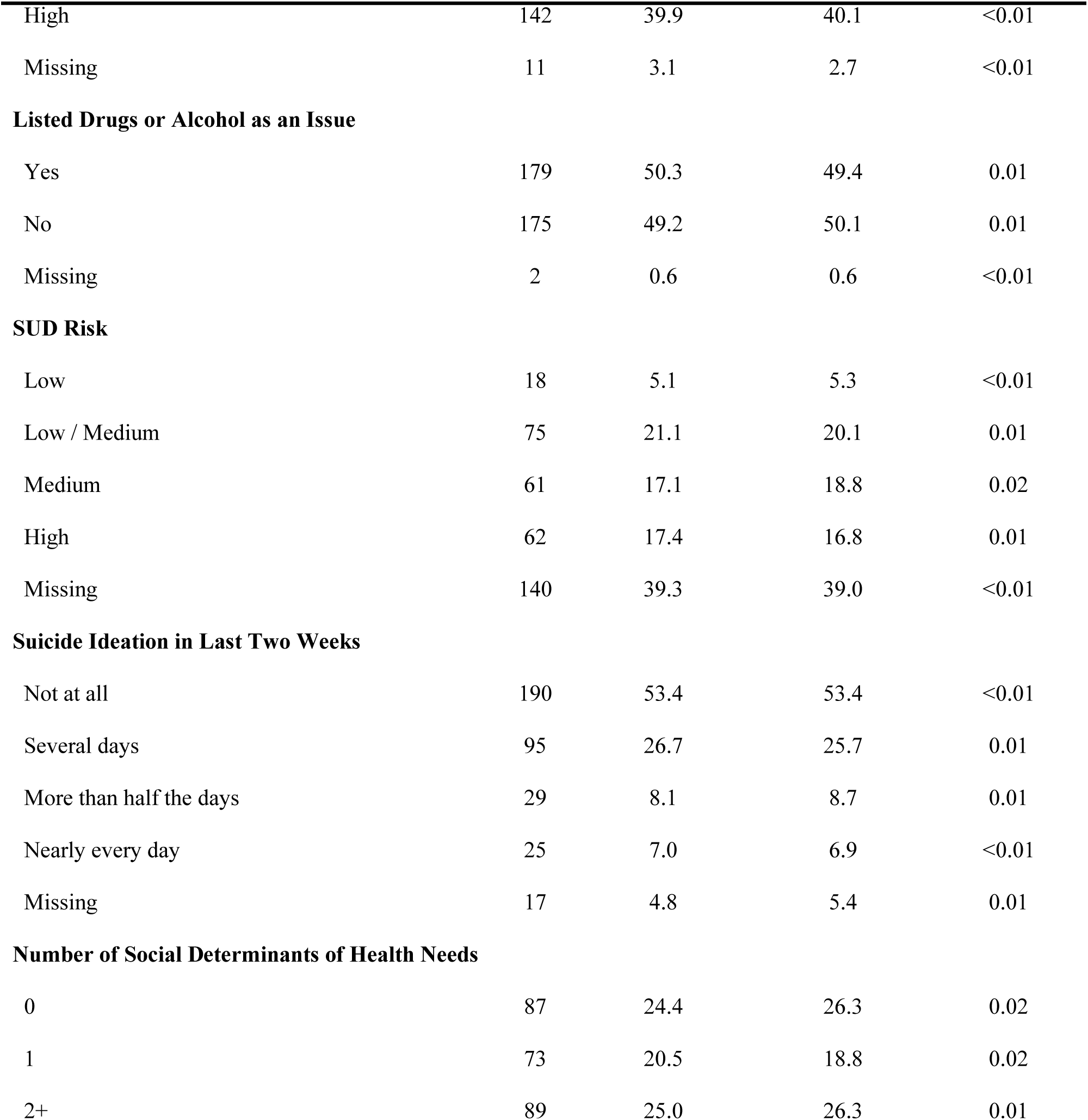
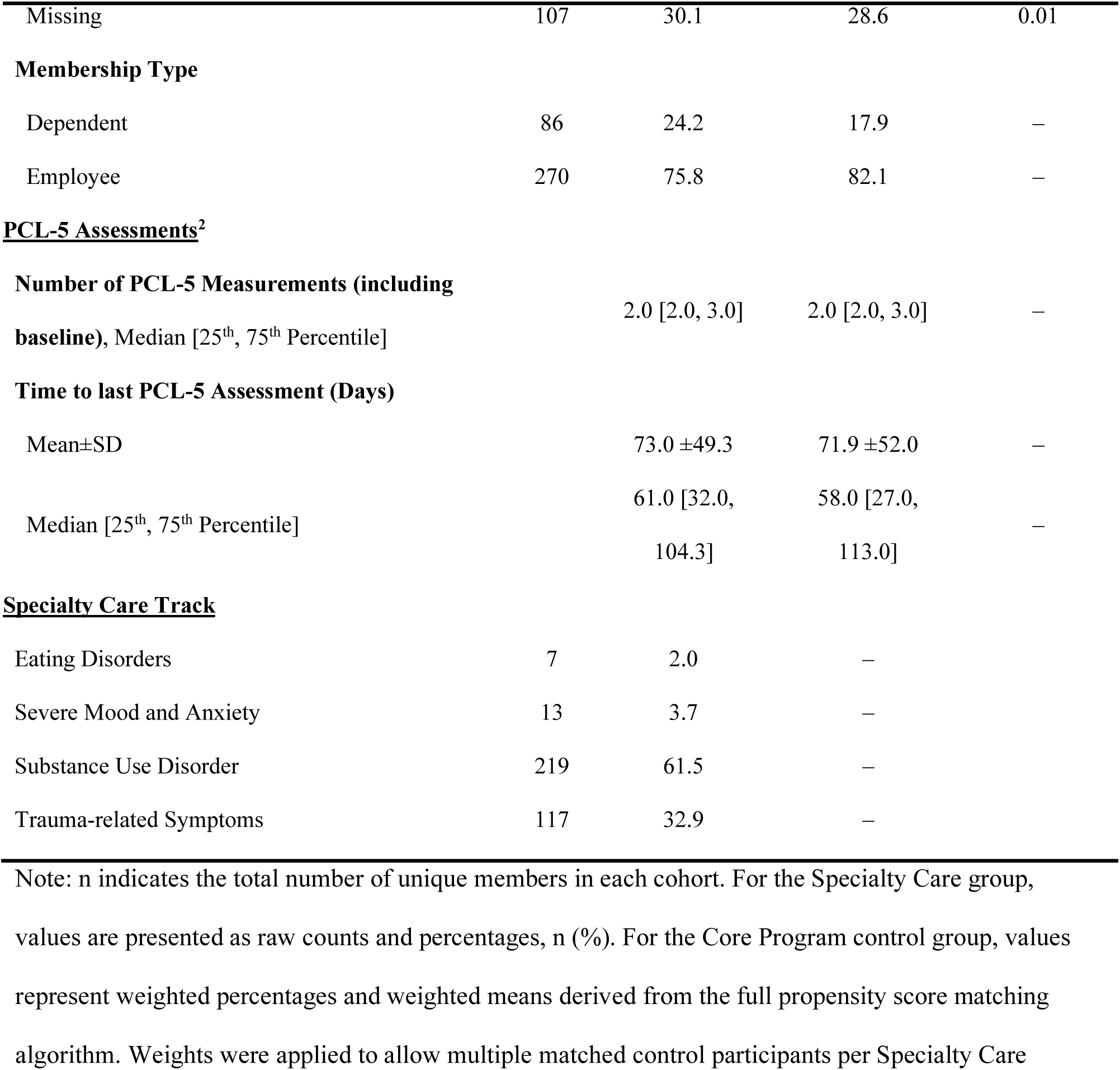
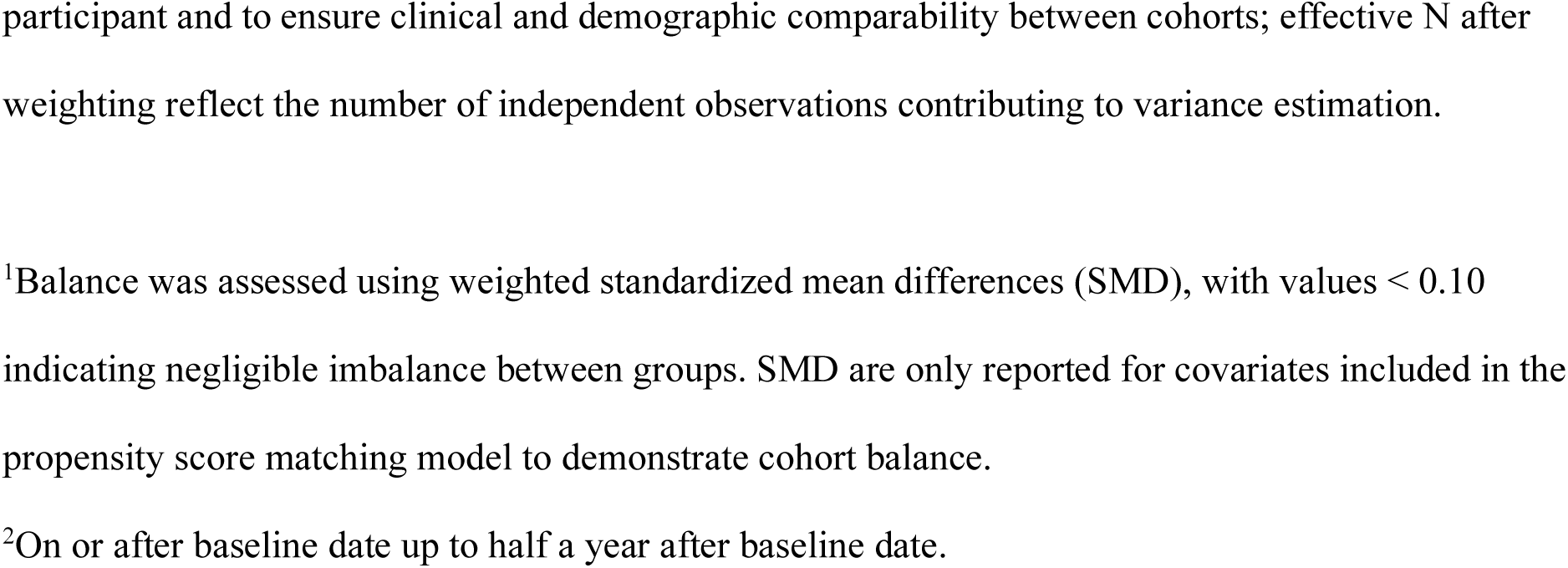
Baseline Characteristics for the PCL-5 Analysis After Matching.

### Psychotherapy Dose and Timing, and Clinician Training

Psychotherapy initiation was high and similar across groups (95% SpC; 94% Core), but SpC participants received 31% more psychotherapy (expected cumulative sessions 6.9 vs 5.3 by 12 weeks) and faster early follow-up (Table 2). By day 14, 77.7% of SpC participants versus 68.7% of Core completed a second session. SpC participants were 40% more likely to complete a second session (Hazard Ratio [HR], 1.4; 95% CI, 1.2–1.7), with higher expected session counts by day 14 (2.1 vs 1.7). SpC participants more frequently engaged in medication management (26% vs 21%; expected session counts among those engaged 2.0 vs 1.7). Care navigation engagement was also higher in SpC (84% vs 1.0%; expected navigation session counts among those engaged 2.9 vs 0.9).

**Table 2.** Clinician Training and Service Utilization During Follow-up.

| Measure | Specialty Care |  | Core Program<br>(weighted) |  |
| --- | --- | --- | --- | --- |
|  | Estimate | 95%CI | Estimate | 95%CI |
| Had $\geq 1$ therapy session during study period, % | 95.4 | | 94.4 | |
| Expected number of therapy sessions<sup>1</sup> | 6.93 | 6.44, 7.42 | 5.28 | 5.09, 5.48 |
| Had $\geq 1$ medication management session during study period, % | 26.3 | | 20.7 | |
| Expected number of medication management sessions<sup>2</sup> | 2.05 | 1.72, 2.37 | 1.68 | 1.51, 1.85 |
| Had $\geq 1$ care navigation appointment during study period, % | 84.2 | | 1.0 | |
| Expected number of care navigation sessions<sup>2</sup> | 2.95 | 2.79, 3.11 | 0.91 | 0.76, 1.06 |
| Had $\geq 1$ psychotherapy session with a clinician with documented trauma-focused training during study period, %<sup>3</sup> | 85.4 | | 79.2 | |

Note: Expected session counts were estimated using weighted mean cumulative functions. Percentage rows show raw percentages for Specialty Care and weighted percentages for Core.
<sup>1</sup>Among those with at least one therapy session within 30 days of baseline. To assess the intensity of therapy, follow-up began on the date of the first therapy session and continued for 12 weeks.
<sup>2</sup>Among those with at least one medication/care navigation session since baseline. Follow-up began at baseline and continued for 12 weeks to capture all post-baseline activity.
<sup>3</sup>Among those with at least one psychotherapy session. Trauma-focused training included Prolonged Exposure, Cognitive Processing Therapy, Eye Movement Desensitization and Reprocessing, Accelerated Resolution Therapy, or trauma-focused Cognitive Behavioral Therapy.

Among participants with at least one psychotherapy session, 85% in SpC vs 79% in Core had at least one session with a clinician with documented trauma-focused training. Within the SpC trauma track, all participants with psychotherapy exposure had at least one session with a clinician with documented trauma-focused training.

### Symptom Trajectories

Both groups demonstrated significant PTSD symptom improvement. Over 16 weeks, PCL-5 scores decreased by an average of 12.8 points in Core (95%CI 12.5, 13.2; *d* =1.01) and 16.6 points in SpC (95%CI 14.7, 18.5; *d* =1.31) (Figure 1). The SpC-by-time interaction was significant (β = −1.3 per log-week, p < .001; Table 3), indicating faster symptom reduction in SpC. The between-group difference in change was 3.8 points (95%CI 1.8, 5.7; *d* =0.30), representing a modest incremental effect. Divergence emerged within the first month and widened over time.

**Figure 1.**
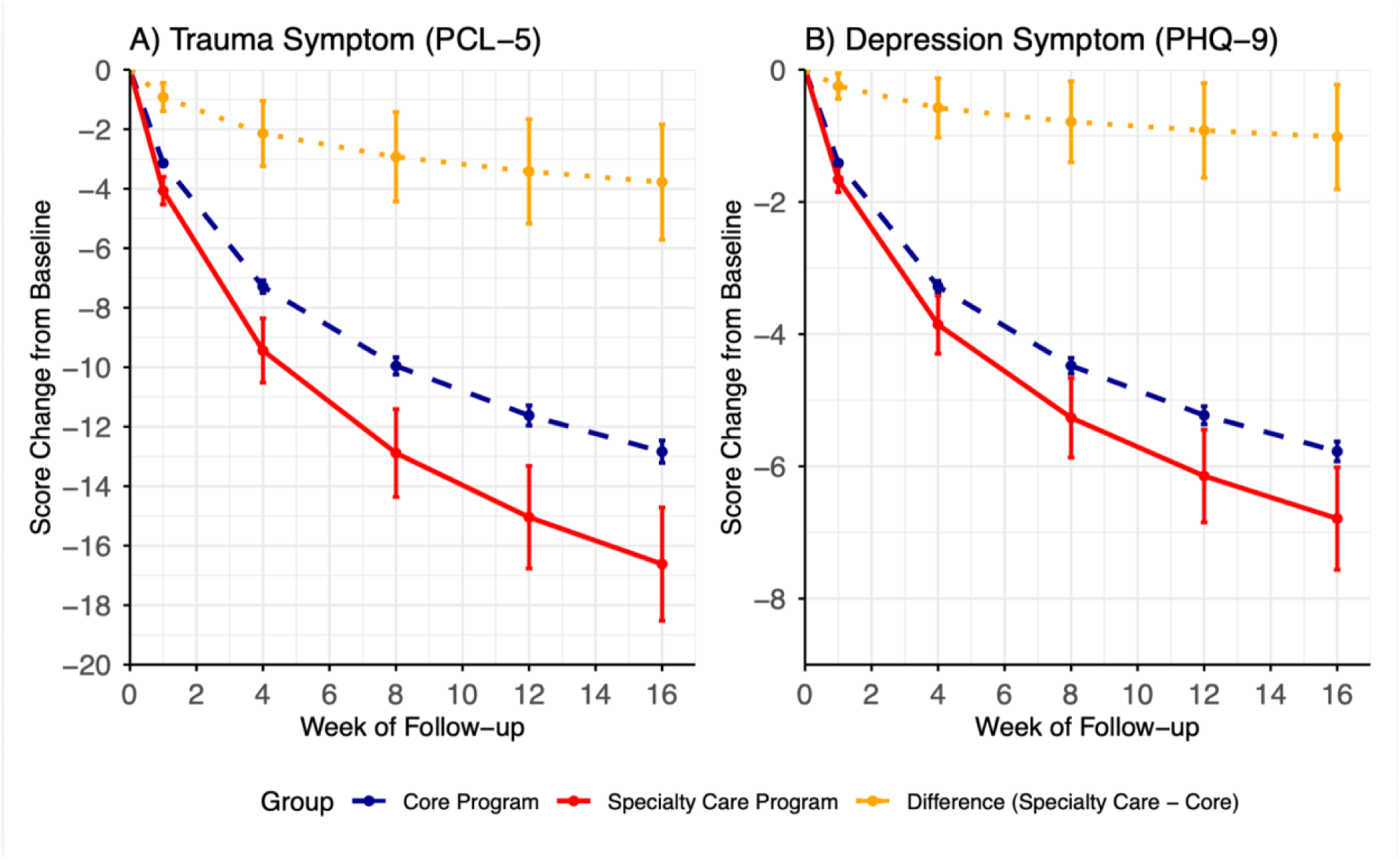
Predicted Symptom Trajectories for PTSD and Depression Over Time. Modeled changes in (A) PCL-5 and (B) PHQ-9 scores based on linear mixed-effects models using log(weeks) from baseline. The red and blue lines represent the predicted mean scores for the Specialty Care and Core Program groups, respectively. The yellow line illustrates the estimated difference in scores between the Specialty Care and Core Program groups over a 16-week period, highlighting the widening gap in symptom improvement. Vertical error bars represent 95% confidence intervals at 1, 4, 8, 12, and 16 weeks. PCL-5, Posttraumatic Stress Disorder Checklist for DSM-5; PHQ-9, Patient Health Questionnaire 9-item Scale Specialty Care showed greater PTSD and depression symptom improvement than Core over 16 weeks.

**Table 3.** Regression Estimates of Change in PTSD and Depression Symptoms.

|  | PTSD (PCL-5 score) |  |  | Depression (PHQ-9 score)<sup>1</sup> |  |  |
| --- | --- | --- | --- | --- | --- | --- |
|  | 27,791 Observations |  |  | 24,855 Observations |  |  |
|  | 9,762 unique patients |  |  | 8,313 unique patients |  |  |
|  | 355 matched groups |  |  | 294 matched groups |  |  |
| **Fixed Effect** | **Estimate<sup>2</sup>** | **95% CI** | **p-value** | **Estimate<sup>3</sup>** | **95% CI** | **p-value** |
| Intercept | 45.6 | 44.7, 46.5 | <0.001 | 16.7 | 16.3, 17.0 | <0.001 |
| Log-weeks | -4.5 | -4.7, -4.4 | <0.001 | -2.0 | -2.1, -2.0 | <0.001 |
| Specialty Care Program | -0.2 | -1.9, 1.5 | 0.8 | -0.03 | -0.7, 0.6 | 0.9 |
| Log-weeks*Specialty Care | -1.33 | -2.0, -0.6 | <0.001 | -0.4 | -0.6, -0.1 | 0.01 |
| **Random effect** | **SD** |  |  | **SD** |  |  |
| Patient ID | 10.9 |  |  | 3.9 |  |  |
| Match ID | 0.7 |  |  | 0.6 |  |  |
| Residual | 11.6 |  |  | 4.4 |  |  |

PCL-5, Posttraumatic Stress Disorder Checklist for DSM-5; PHQ-9, Patient Health Questionnaire 9-item Scale; CI, Confidence Interval; SD, Standard Deviation
<sup>1</sup>Among those with baseline PHQ-9 $\geq 10$
<sup>2</sup>The linear mixed-effects model was estimated using restricted maximum likelihood. Fixed effects included the logarithm of weeks since baseline, treatment group (Specialty Care Program vs. Core Program), and their interaction, with random intercepts for patient ID and Match ID. Baseline PHQ-9 scores ( $<15$ or $\geq 15$ ) and high risk of substance use disorder were included as covariates to adjust for baseline clinical severity.
<sup>3</sup>The linear mixed-effects model was estimated using restricted maximum likelihood. Fixed effects included the logarithm of weeks since baseline, treatment group, and their interaction, with random
intercepts for patient ID and Match ID. High risk of substance use disorder was included as a clinical covariate.

PHQ-9 decreased by 5.8 points in Core over 16 weeks (95%CI 5.6, 5.9; *d* =1.23) and 6.8 points in SpC (95%CI 6.0, 7.6; *d* =1.44). The SpC-by-time interaction was small but significant (β = −0.4 per log-week, p =.01), with a 1.0-point between-group difference (95%CI 0.2, 1.8; *d* =0.22).

### Clinical Milestones

At any given time point, SpC participants had a 31% higher hazard of symptom-defined PTSD recovery (PCL-5 < 31) compared to Core (HR 1.31; 95%CI: 1.10, 1.57; Table 4). Similar associations were observed for low-symptom status (PCL-5 ≤20; HR 1.32, 95%CI: 1.03, 1.68), clinically meaningful improvement (≥10-point drop; HR 1.16; 95%CI: 1.01, 1.34), and the composite recovery plus clinically meaningful improvement (HR 1.24; 95%CI: 1.03, 1.51). At 12 weeks, predicted recovery was 29% (SpC) versus 23% (Core); low-symptom status 9% vs. 7%; and clinically meaningful improvement 58% vs 53% (eTable 2). SpC was not significantly associated with faster PHQ-9 milestones (recovery 38% vs 35% at 12 weeks).

**Table 4.** Hazard Ratios for Time to Symptom-based Recovery, Low-symptom Status, and Clinically Meaningful Improvement.

| Time to Event | PTSD |  |  | Depression |  |  |
| --- | --- | --- | --- | --- | --- | --- |
|  | HR | 95%CI | p-value | HR | 95%CI | p-value |
| Recovery | 1.31 | 1.10, 1.57 | 0.003 | 1.10 | 0.91, 1.31 | 0.37 |
| Low-symptom Status | 1.32 | 1.03, 1.68 | 0.02 | 1.37 | 0.99, 1.88 | 0.05 |
| Clinically Meaningful Improvement | 1.16 | 1.01, 1.34 | 0.04 | 1.06 | 0.90, 1.24 | 0.50 |
| Clinically Meaningful Improvement & Recovery | 1.24 | 1.03, 1.51 | 0.02 | 1.08 | 0.88, 1.33 | 0.48 |

HR, Hazard Ratio; CI, Confidence Interval

### Sensitivity and Subgroup Analysis

Findings were consistent across sensitivity analyses (eFigures 3–5). The association between SpC and faster PTSD recovery was strongest in the trauma pathway (HR 1.68; 95% CI, 1.31–2.16) and positive but non-significant in the SUD pathway (HR 1.09; 95% CI, 0.87–1.36), indicating that the overall estimate is driven substantially by the trauma subgroup despite SUD pathway comprising the majority of SpC participants. Psychotherapy dose also differed markedly by track: the trauma track received nearly twice as many adjusted expected sessions by 12 weeks as the SUD track (9.58 vs 5.41), and SUD-track dose was only marginally higher than the Core comparator (5.3) (eTable 3).

### Post hoc Analysis

Adjusting for cumulative psychotherapy exposure attenuated the SpC-by-time interaction to non-significance, suggesting that faster symptom reduction in SpC was largely attributable to greater therapy dose rather than differential per-session effectiveness. Greater cumulative therapy exposure was strongly associated with lower PTSD severity, with larger marginal effects earlier in treatment (interaction p < .001; eTable 4). Additional adjustment for care navigation exposure did not materially change these associations, and navigation was not independently associated with symptom change.

## Discussion

In this matched cohort of adults with elevated PTSD symptoms receiving routine outpatient care, SpC participation was associated with modestly faster symptom-defined recovery than standard outpatient care. The most parsimonious interpretation, supported by our post hoc analyses, is that the model worked primarily by delivering more psychotherapy sooner rather than by making each session more effective. Care navigation may have been a key implementation mechanism by facilitating dose and engagement, while documented trauma-focused clinician training may have supported appropriate PTSD care, particularly in the trauma pathway. This is consistent with evidence that frequent, front-loaded sessions can match or exceed distributed treatment,^7,9,11^ plausibly by counteracting avoidance-driven disengagement early in care when proactive scheduling and outreach matter most.

This interpretation is reinforced by a dose gradient across tracks: the trauma track received the most psychotherapy and showed the strongest recovery association (HR 1.68), whereas the SUD track received a dose close to Core and showed no significant advantage in PTSD outcomes (HR 1.09). This gradient is consistent with a dose mechanism, but may also reflect differences in clinical complexity and treatment sequencing. Because hierarchical triage prioritized substance-use stabilization and the trauma track excluded comorbid SUD, these findings should therefore not be interpreted as evidence that patients with co-occurring PTSD and SUD symptoms should be routed preferentially to trauma-focused care. Rather, they raise an implementation question about when such patients may benefit from earlier trauma-focused assessment, integrated PTSD/SUD treatment planning, or transition from stabilization to trauma-focused care.^20^ Overall symptom improvement across SpC and the positive but nonsignificant association in the SUD pathway are consistent with evidence that high-quality SUD treatment can yield PTSD symptom improvement even when trauma is not the focus.^21^

Although SpC was associated with steeper depressive-symptom decline, this did not translate into faster categorical depression milestones, possibly reflecting the established effectiveness of the Core program for depression^22^ and limited power in the depression-eligible cohort.

For health systems and payers, these findings suggest that PTSD symptom outcomes may improve by re-engineering aspects of outpatient delivery rather than introducing novel treatments. Practical targets include rapid second-session follow-up, higher early session density, measurement-based care, proactive outreach to promote treatment initiation, and ongoing navigation to support engagement. Because the clearest measured pathway was earlier and greater psychotherapy exposure, targeted operational changes, such as reserved early-follow-up slots, automated scheduling, and outreach to patients at risk for dropout, could be tested incrementally within existing benefit structures.

Feasibility, staffing, and cost remain central questions. Higher session density and proactive navigation require additional capacity and likely carry incremental cost. Lacking cost or utilization-offset data, we cannot address cost-effectiveness or reduced reliance on intensive services.

This study has several limitations. First, despite matching, the retrospective design cannot eliminate employer- or individual-level unobserved confounding. Second, trauma exposure and clinician-diagnosed PTSD were unavailable; participants were identified by symptom threshold, and recovery outcomes are symptom-defined proxies. Third, baseline definitions differed between groups; anchoring SpC baseline to enrollment may have missed pre-enrollment improvement, likely biasing toward the null. Fourth, treatment fidelity and asynchronous engagement were unmeasured and may have contributed to observed differences. Finally, the sample comprised employed, benefit-eligible adults using a digital platform with relatively high engagement and follow-up data and may not generalize to populations with greater social-determinants barriers, uninsured populations, or non-digital settings.

## Conclusions

In an employer-sponsored digital mental health platform, a structured outpatient delivery model incorporating condition-focused clinicians, higher session density, and proactive navigation was associated with modestly faster symptom-defined PTSD recovery, an effect that appeared to be driven largely by greater and earlier psychotherapy dose. These findings suggest that redesigning the structure and timing of care delivery may be a practical lever for improving routine PTSD outcomes without requiring a novel psychotherapy protocol.

## Disclosures and Acknowledgments

Drs. Khor, Klempner, Dworkin, Schwehm, Brown, Chekroud, and Hawrilenko report being employed by and holding equity in Spring Care Inc. Dr. Chekroud reports being the lead inventor on three patent submissions relating to treatment for major depressive disorder: U.S. Patent and Trademark Office number Y0087.70116US00 and provisional application numbers 62/491660 and 62/629041. Dr. Dworkin reports receiving consulting fees from Flourish Science. Dr. Chekroud also reports holding equity in Carbon Health Technologies Inc., Wheel Health Inc., Parallel Technologies Inc., Healthie Inc., and UnitedHealthcare; receiving consulting fees from Fortress; and providing unpaid advisory services to health care technology startups. No support from pharmaceutical companies was received. The authors report no additional acknowledgments or disclosures.

## Previous Presentation

These data have not been previously presented.

## Data Availability

The datasets generated or analyzed during this study are not publicly available because they contain sensitive personal health information. Aggregate findings may be available from the corresponding author upon reasonable request.

