## Supplemental Material for "Structured Outpatient Specialty Care and Faster Posttraumatic Stress Disorder Symptom Improvement: A Matched Cohort Study"

### Appendix

#### eTable 1. Core Program vs. Specialty Care Program Design

| Component | Core Program | Specialty Care Pathway | Measurement in Current Analysis |
|---|---|---|---|
| **Benefit access** | Available to all members. | Available only when offered by employer benefit design. | Program access and enrollment observed; employer-level reasons for benefit selection were not measured. |
| **Clinician training** | Licensed outpatient clinicians were not required to have training in trauma-focused therapies; members could self-select clinicians with documented trauma-focused training. | Licensed clinicians with condition-focused training and/or certification in manualized protocols. All trauma-pathway clinicians were required to have trauma-focused training, defined as training in at least one of Prolonged Exposure, Cognitive Processing Therapy, Eye Movement Desensitization and Reprocessing, Accelerated Resolution Therapy, or trauma-focused Cognitive Behavioral Therapy. For complex PTSD presentations, clinicians could provide an initial three-session stabilization phase informed by Skills Training in Affective and Interpersonal Regulation prior to initiating PTSD-focused treatments. | Clinician assignment and clinician training on trauma-focused protocols were observed. |
| **Scheduling Appointments** | Patients predominantly self-scheduled therapy appointments. | Condition-specific navigators provided proactive outreach, including an approximately 45-min triage intake, to facilitate scheduling and therapy initiation. | Timing of psychotherapy initiation and early follow-up were measured. |
| **Psychotherapy** | 55-minute outpatient psychotherapy. | Higher-density 55-minute outpatient psychotherapy, up to twice-weekly when clinically indicated. | Session frequency and early session density were measured; actual protocol selection, session content, fidelity, and protocol completion were not observed. |
| **Care navigation** | Access facilitation and coordination; contacts were typically brief and front-loaded around intake. | Ongoing follow-up 1–2 times per month by condition-specific navigators to monitor acuity, support engagement, reduce early termination, and escalate care when indicated. | Navigation visit occurrence and volume were measured; outreach attempts and content were not fully measured. |
| **Medication Management** | Available as needed | | Medication-management visit occurrence and cumulative visit count were measured; medication prescribing details were not measured. |

| | | |
|---|---|---|
| **Digital care infrastructure and asynchronous engagement** | Scheduling, telehealth, messaging, symptom assessments, self-guided tools, and structured between-session engagement (e.g., practice assignments, behavioral experiments, review of session recordings, psychoeducation, and digital tools). | Specific feature use and between-session utilization not analyzed. |

**eFigure 1. Consort Diagram**

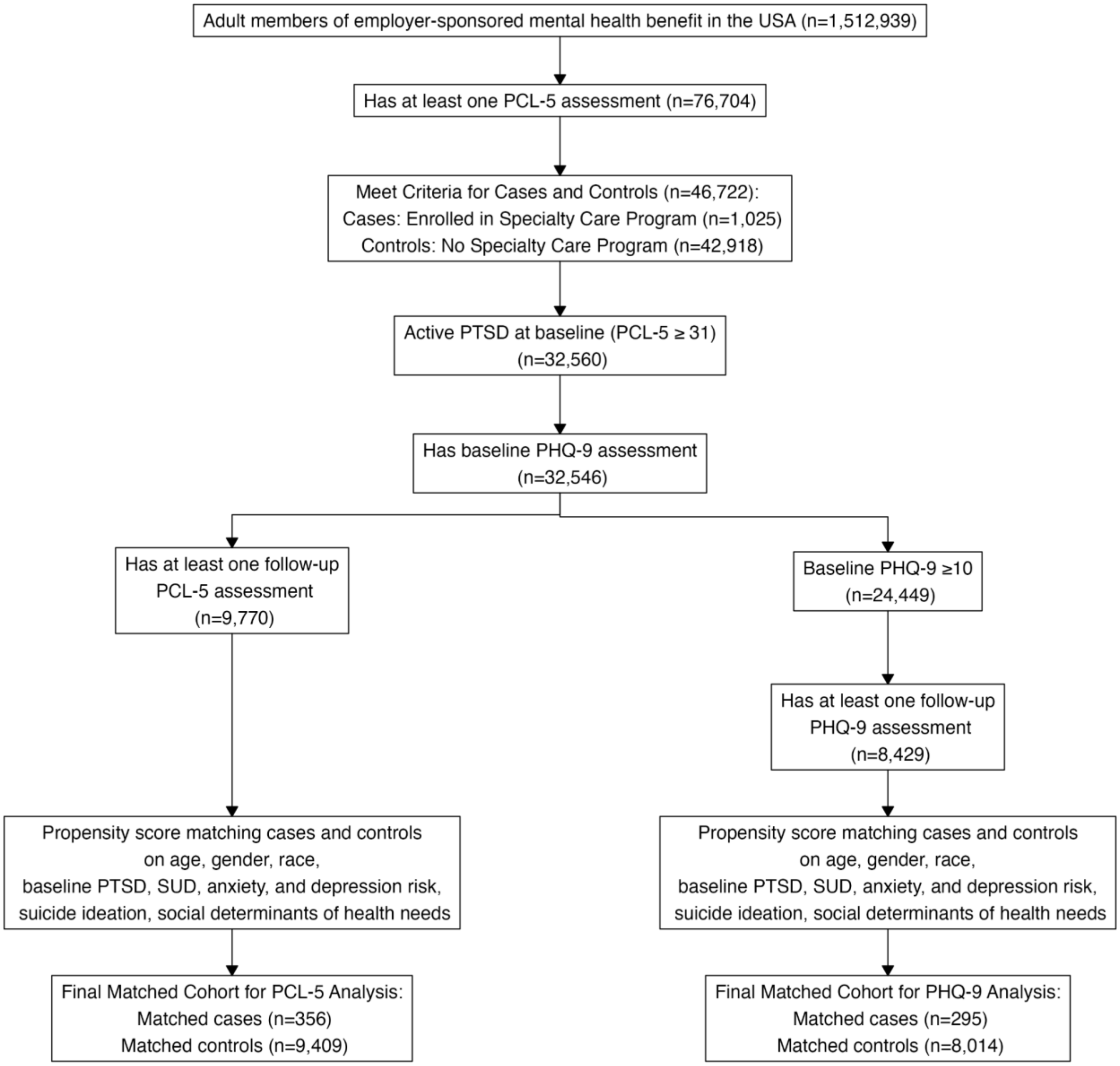

**eText 1. Analysis of Selective Attrition and Baseline Comparability**
To evaluate potential attrition bias, baseline characteristics were compared between participants with at least one follow-up PCL-5 assessment and those with baseline PCL-5 only. Groups were comparable across demographics and clinical severity. Participants with no follow-up had slightly higher prevalence of 2 or more social determinants of health needs at intake, suggesting attrition was not strongly selective with respect to clinical severity, although social instability may have affected longitudinal assessment completion.

**Baseline characteristics by Follow-up Status (before matching)**

| | **No follow-up (n=22,776)** | **≥ 1 follow-up (n=9,770)** |
|---|---|---|
| **Age (Years),** Mean (SD) | 34.7 (10.7) | 36.4 (11.2) |
| **Woman** | 13775 (60.5%) | 6132 (62.8%) |
| **Race and Ethnicity** | | |
| Black or African American | 1649 (7.2%) | 673 (6.9%) |
| Hispanic or Latino | 752 (3.3%) | 310 (3.2%) |
| Non-Hispanic White | 3319 (14.6%) | 1413 (14.5%) |
| **PCL-5 Score,** Mean (SD) | 51.5 (12.7) | 51.5 (12.7) |
| **PHQ-9 Score**, Mean (SD) | 14.4 (6.30) | 14.5 (6.32) |
| **Medium or High Anxiety Level (GAD-7)** | 10804 (47.4%) | 4821 (47.3%) |
| **Listed Drugs or Alcohol as an Issue** | 2393 (10.5%) | 969 (9.9%) |
| **High SUD Risk** | 845 (3.7%) | 327 (3.3%) |
| **Suicide Ideation ≥ Several Days in Last Two Weeks** | 8,077 (35.5%) | 3369 (34.5%) |
| **2+ Social Determinants of Health Needs** | 6160 (27.0%) | 2421 (24.8%) |

**eFigure 2: Covariate Balance After Propensity Score Matching**

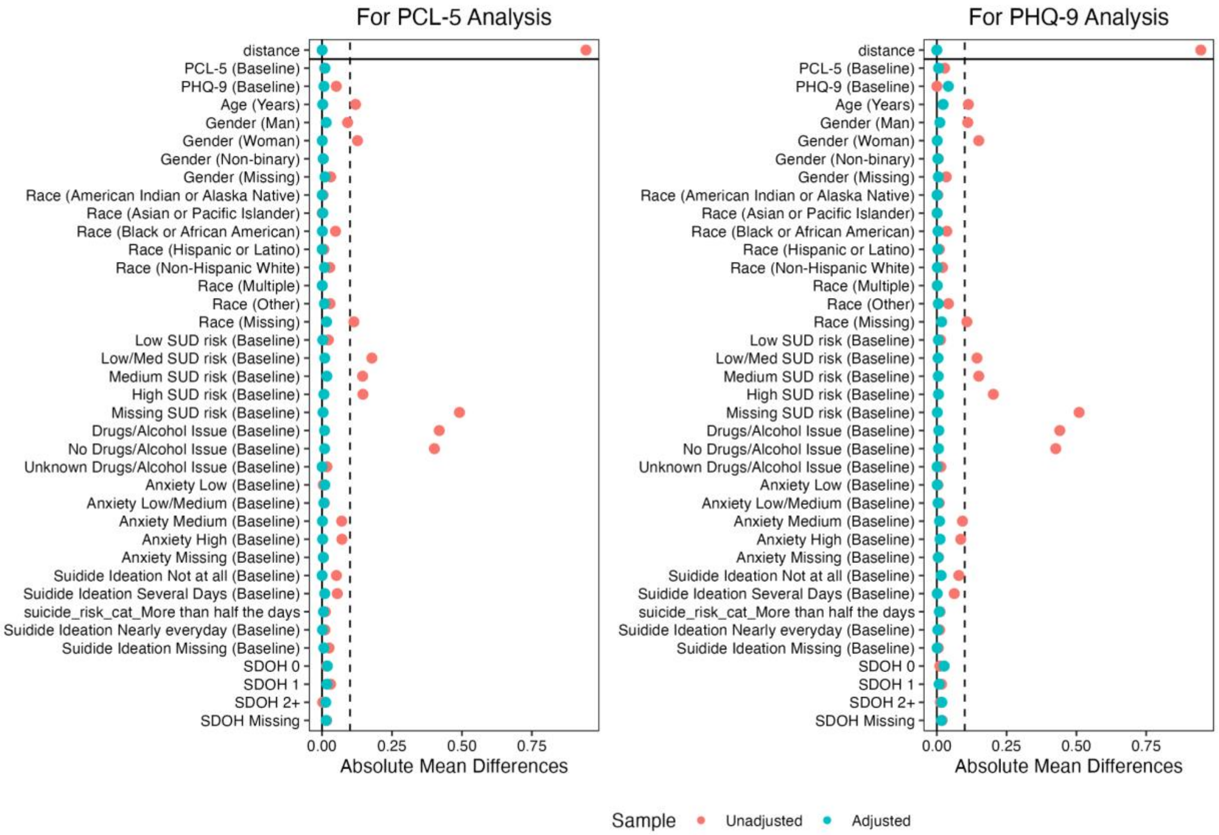


Note: all SMD for both analyses are <0.10 post-matching.

**eTable 2. Predicted Probability of Symptom-defined Recovery, Low-symptom Status, and Clinically Meaningful Improvement at 12 weeks**

| | PTSD, % (95% CI) | | | Depression, % (95% CI) | | |
|---|---|---|---|---|---|---|
| | **Recovery** | **Low-symptom Status** | **Clinically Meaningful Improvement** | **Recovery** | **Low-symptom Status** | **Clinically Meaningful Improvement** |
| **Program** | PCL-5<31 | PCL-5≤20 | PCL-5 change 10+ points | PHQ-9<10 | PHQ-9<5 | PHQ-9 change 5+ points |
| Specialty Care | 28.7% (24.3, 33.1) | 9.4% (7.1, 11.8) | 58.0% (53.0, 63.1) | 37.7% (32.3, 43.1) | 6.7% (4.6, 8.8) | 58.4% (52.6, 64.1) |
| Core Program | 22.7% (21.3, 24.1) | 7.2% (6.4, 8.1) | 52.6% (51.3, 53.8) | 35.2% (33.8, 36.7) | 4.9% (4.3, 5.6) | 56.4% (55.0, 57.8) |

**eFigure 3. Sensitivity Analysis of Predicted PTSD and Depression Symptom Trajectories.** Vertical error bars represent 95% confidence intervals at 1, 4, 8, 12, and 16 weeks.

**<u>i) Only those engaged in care, defined as starting therapy within 30 days of baseline date</u>**

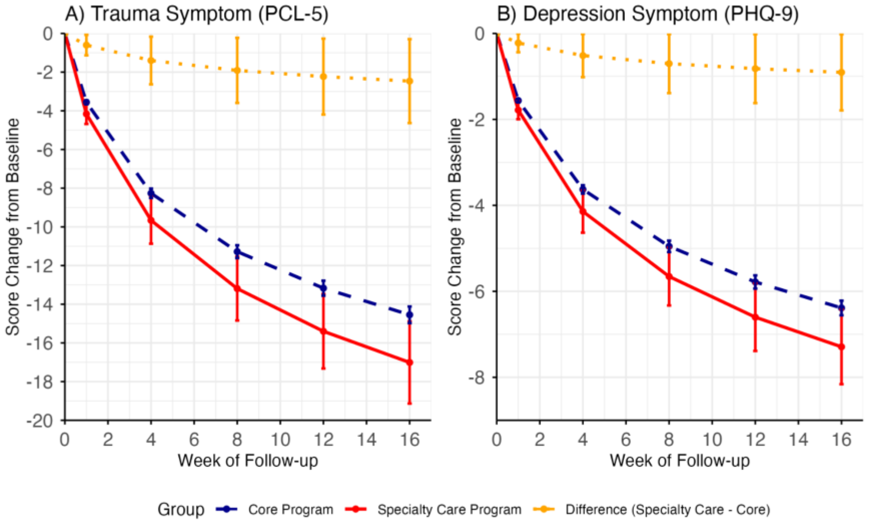


**<u>ii) Trauma-Care Pathway Subgroup</u>**

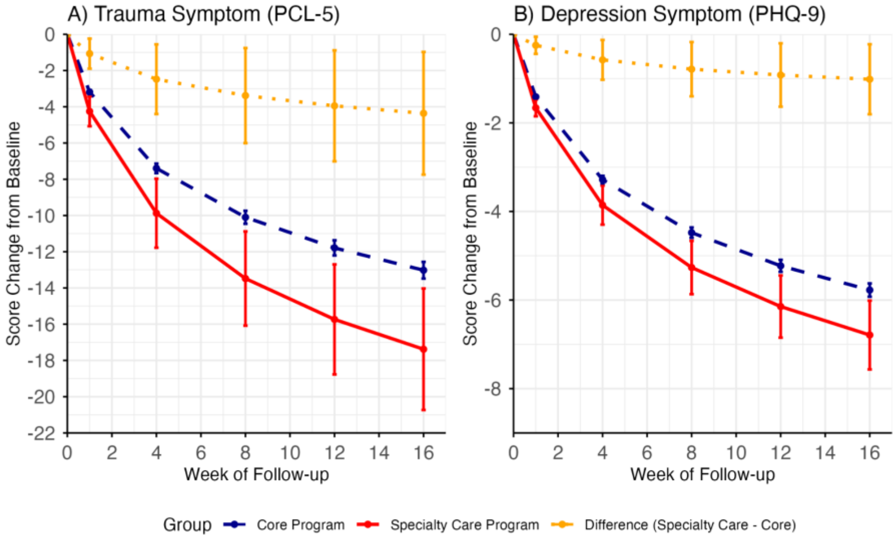

**iii) SUD-Care Pathway Subgroup**

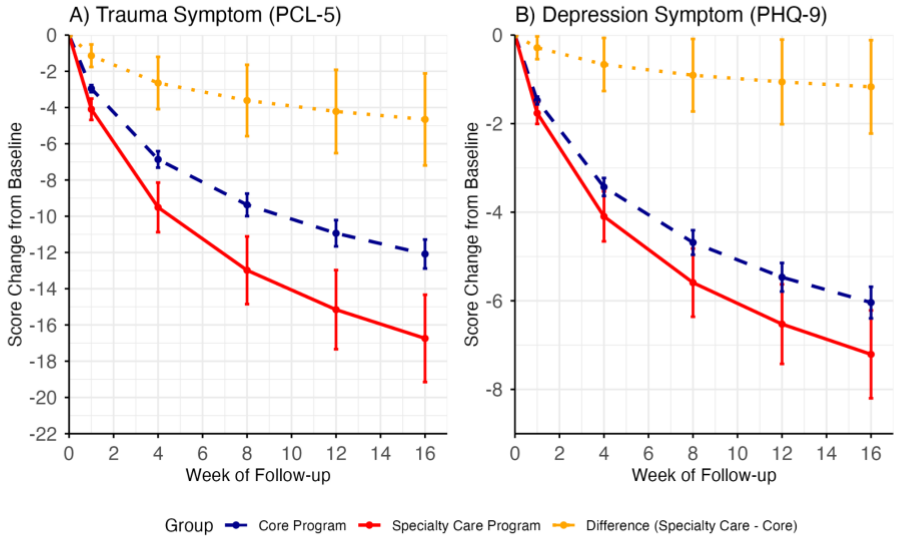

**eFigure 4. Sensitivity Analysis of PTSD (A) and Depression (B) Symptom Trajectories: Interaction Effect of Specialty Care and Time.**

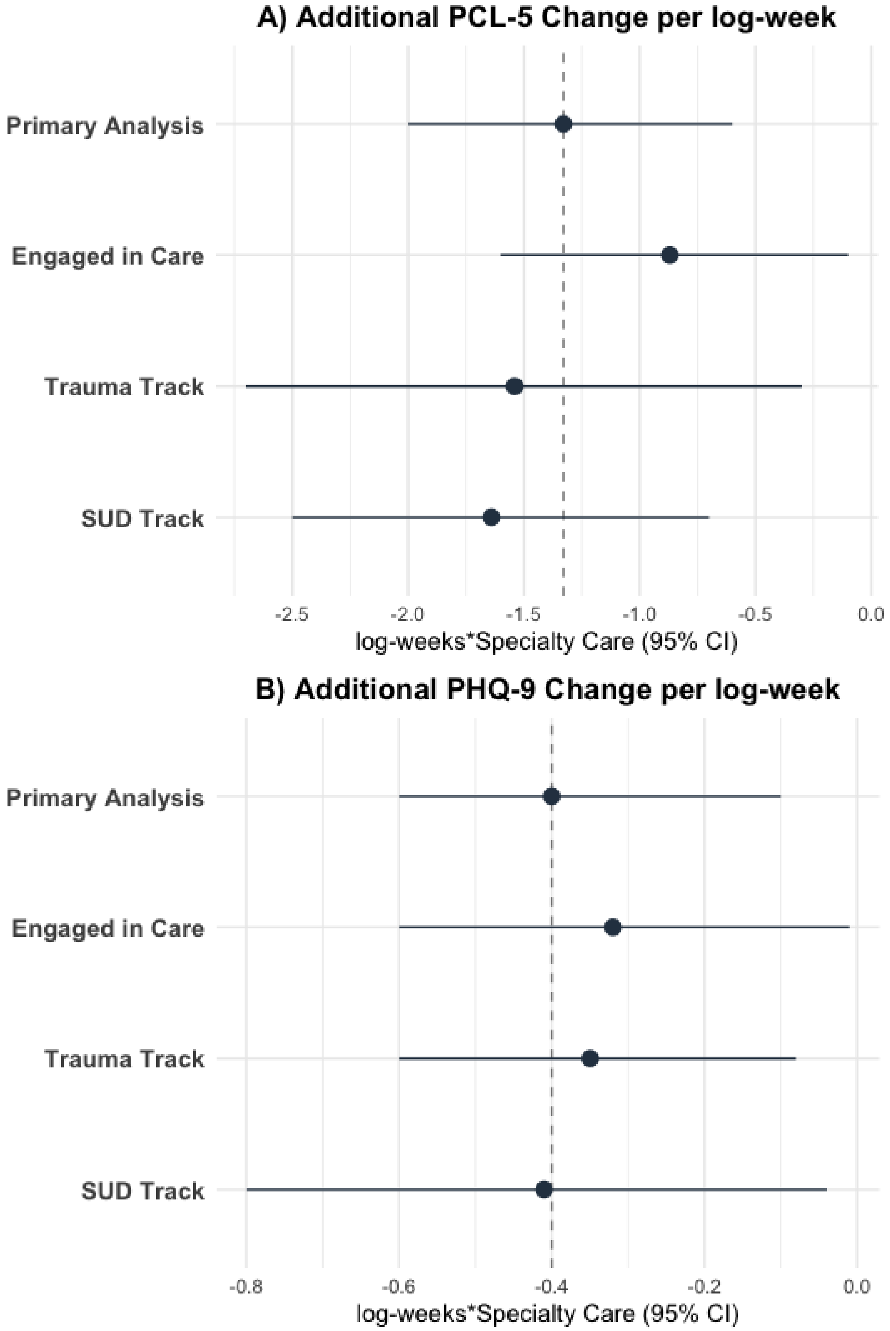


Note: Estimates represent the beta coefficients for the interaction term (Specialty Care*log-week) from the Linear Mixed-Effects Models. These values reflect the incremental rate of symptom reduction in Specialty Care relative to matched Core Program.

**eFigure 5. Sensitivity Analyses - Hazard Ratios for Time to Recovery**

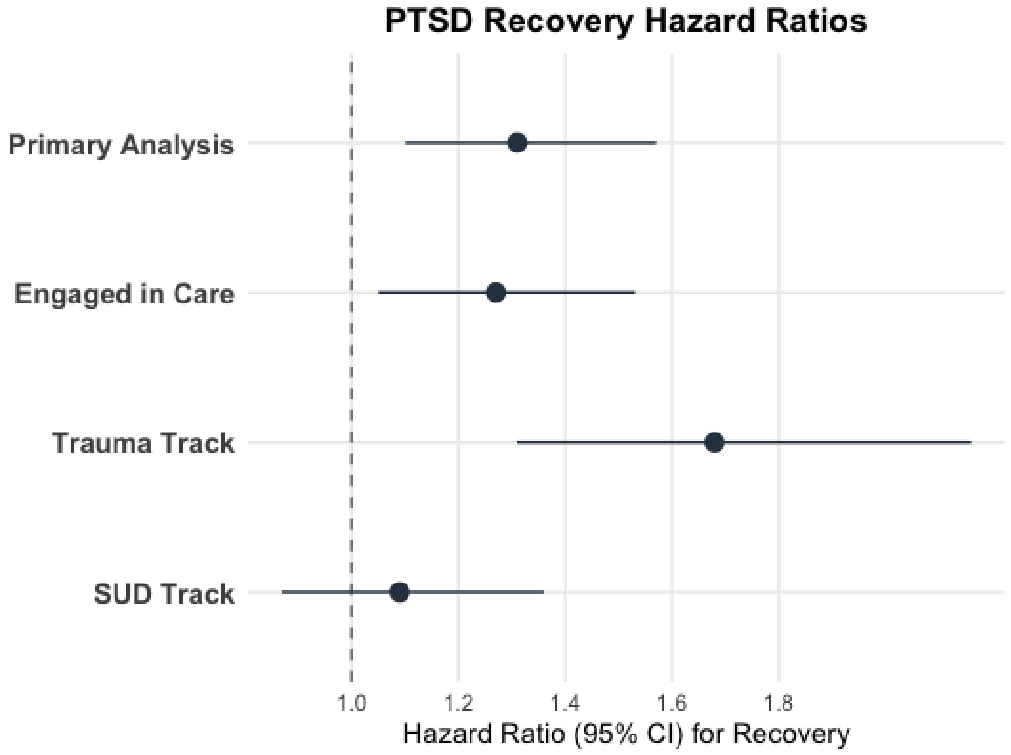

**eTable 3. Sensitivity Analyses - Expected service utilization at 12 weeks in the trauma vs. SUD track, estimated using weighted mean cumulative function**

| Measure | Trauma Track | SUD Track |
|---|---|---|
| | Expected mean (95% CI) | Expected mean (95% CI) |
| **Expected # of therapy sessions**[1] | 9.58 (8.54, 10.6) | 5.41 (5.02, 5.80) |
| **Expected # of medication management sessions**[2] | 2.49 (1.45, 3.52) | 1.89 (1.59, 2.18) |
| **Expected # of care navigation sessions**[2] | 3.77 (3.51, 4.03) | 2.54 (2.36, 2.72) |

**eTable 4. Post Hoc Mechanistic Analyses of PTSD Symptom Trajectories.** Cumulative therapy and navigation exposure were defined as sessions completed prior to each PCL-5 assessment (same-day sessions assigned to the subsequent interval to preserve temporal ordering). Analyses were restricted to psychotherapy initiators (≤30 days from baseline). Sequential models extended the primary specification by adding (1) cumulative psychotherapy exposure and interaction terms and (2) cumulative navigation exposure and interaction. **(**21909 Observations, 7,591 patients, 289 matched groups.)

| Fixed Effect[1] | Base Model | Therapy-adjusted | Therapy- and Navigation-adjusted |
|---|---|---|---|
| **Intercept** | 45.4*** | 46.0*** | 46.0*** |
| **Log-weeks** | -5.1*** | -4.0*** | -4.0*** |
| **Specialty Care Program** | -0.6 | -0.5 | -0.5 |
| **Log-weeks*Specialty Care** | -0.87* | -0.78 | -0.84 |
| **Cumulative Therapy** | | -2.6*** | -2.6*** |
| **Log-weeks*therapy** | | 0.77*** | 0.77*** |
| **Specialty Care*therapy** | | 0.1 | 0.1 |
| **Cumulative Navigation** | | | 0.2 |
| **Log-weeks*navigation** | | | -0.07 |

SD, Standard Deviation; * p < .05 ** p < .01 *** p < .001

[1]Random intercepts included for participants and matched strata.
